# Effectiveness of 20-valent pneumococcal conjugate vaccine against pneumococcal pneumonia hospitalizations among U.S. adults: Test-Negative Design

**DOI:** 10.64898/2026.09.13.26362937

**Authors:** A J Ologunowa, S Kogut, V L Vrishali, SA Cohen, A R Caffrey

**Author notes:** **Corresponding Author:** Aisling R. Caffrey, PhD, MS Professor, Health Outcomes, Department of Pharmacy Practice and Clinical Research, College of Pharmacy, University of Rhode Island, Kingston, RI, USA. 02881, **Co-corresponding Author:** Abiodun J. Ologunowa, MS, BPharm Ph.D. Candidate, Health Outcomes, Department of Pharmacy Practice and Clinical Research, College of Pharmacy, University of Rhode Island Kingston, RI, USA. 02881.

## Abstract

**Objective:** This study estimated the vaccine effectiveness (VE) of 20-valent pneumococcal conjugate vaccine (PCV20) against culture-confirmed pneumococcal pneumonia hospitalizations among adults using a test-negative case-control design.

**Methods:** We included adults (≥18 years) hospitalized with pneumonia in the U.S. Veterans Affairs Healthcare System between January 2023 and December 2025 who had respiratory cultures collected. VE was estimated using multivariable logistic regression adjusted for demographic and clinical characteristics. Subgroup analyses were conducted by age, risk group, and U.S. census region. Sensitivity analyses included inverse probability of treatment weighting (IPTW) and evaluation allowing previous PCV13 vaccination.

**Results:** Among 34,009 pneumonia hospitalizations, 837 (2.5%) were *Streptococcus pneumoniae* culture-positive and 7,428 (21.8%) individuals had received PCV20. PCV20 vaccination was associated with lower odds of culture-confirmed pneumococcal pneumonia hospitalization among adults aged ≥65 years (VE 20.4%; 95% CI: 2.1%, 35.3%), with greater effectiveness observed among those with chronic medical conditions (VE 24.4%; 95% CI: 0.7%, 42.4%). VE was highest in the Western region among adults aged ≥65 years (39.6%; 95% CI: 3.2%, 62.3%), a region with a high burden of serotype 4 disease. Overall, VE among adults aged ≥18 years was 8.8% (95% CI: −8.8%, 23.6%). Findings were consistent across multivariable and IPTW-adjusted analyses.

**Conclusion:** PCV20 provided significant protection against pneumococcal pneumonia hospitalizations among older adults, with greater effectiveness in regions where vaccine-covered serotypes are more prevalent. These findings support the importance of aligning pneumococcal vaccination strategies with regional serotype distribution to maximize vaccine impact.

**Key Summary:** This study provides the first real-world evidence that PCV20 reduces culture-confirmed pneumococcal pneumonia hospitalizations among adults aged ≥65 years, supporting the effectiveness of routine vaccination, with the greatest benefit observed in the Western U.S., where serotype 4 disease is predominant.

## Background

*Streptococcus pneumoniae* remains an important cause of bacterial pneumonia and invasive bacterial disease, especially among older adults and those with underlying conditions.^1^ Collectively, these infections place a considerable burden on patients and the healthcare system, driving pneumonia-related morbidity, mortality, and resource use in the United States (U.S.).^2–4^ About 20%-30% of healthy adults are asymptomatic carriers of *Streptococcus pneumoniae*, creating a substantial reservoir for transmission, particularly for vulnerable populations.^4^ Despite advances in prevention and treatment, pneumococcal pneumonia still poses a major public health challenge. The U.S. Centers for Disease Control and Prevention (CDC) estimates that pneumococcal pneumonia accounts for approximately 150,000 hospitalizations annually and is a frequent complication of influenza infection.^5^ *S. pneumoniae* is associated with about 10-30% of community-acquired pneumonia cases among adults, highlighting its significant impact on respiratory health among adults and is one of the leading causes of death due to infectious diseases in the U.S.^5,6^

The introduction of pneumococcal vaccines has markedly reduced disease caused by vaccine-covered serotypes.^7^ However, widespread pneumococcal conjugate vaccine (PCV) use has reshaped pneumococcal epidemiology through serotype replacement, increasing the importance of broader-valency vaccines and continued evaluation of vaccine effectiveness in real-world settings.^1,4^ CDC Active Bacterial Core surveillance data have demonstrated a disproportionate burden of serotype 4 disease in the Western U.S, with this serotype accounting for more than 30% of invasive pneumococcal disease (IPD) in some settings in that region.^8^ PCV20 was approved by the U.S. Food and Drug Administration (FDA) based on clinical trials demonstrating immunogenicity non-inferior to PCV13 for shared serotypes including serotype 4, and robust immune responses to seven additional serotypes not covered by PCV13.^9,10^ These additional serotypes are estimated to account for approximately 32% of IPD cases among adults aged 19-64 years and 28% among those aged ≥65 years.^10^

Real-world evidence on PCV20 effectiveness remains limited. Although a recent Medicare cohort study evaluated effectiveness against all-cause pneumonia hospitalizations,^11^ the effectiveness of PCV20 against culture-confirmed pneumococcal pneumonia has not been assessed. We therefore conducted a test-negative case-control study to estimate PCV20 effectiveness against culture-confirmed pneumococcal pneumonia hospitalizations in the largest integrated healthcare system in the U.S. and evaluated the geographic variation in vaccine effectiveness across U.S. regions.^2,12^

## Methods

### Study Design, Data Source

We conducted a test-negative case-control study using electronic healthcare data from the U.S. Veterans Affairs (VA) Healthcare System. The VA data systems integrate vaccine administration records, including vaccinations received outside the VA, alongside comprehensive medical history, from inpatient hospitalizations and outpatient visits.^13^

### Study Population and Setting

The study period was January 1, 2023, through December 31, 2025, to capture real-world PCV20 uptake following FDA approval in June 2021 and the subsequent ACIP recommendations update from October 20, 2021.^14^ Eligible patients were VA-active (defined as at least one VA visit in the preceding 12 months) adults (aged ≥18 years) who were hospitalized with pneumonia, identified using primary or secondary International Classification of Diseases, 10th Revision, Clinical Modification (ICD-10-CM) diagnosis codes (Supplementary Table 1),^15^ had a respiratory culture collected on the day prior or during the admission, and had an indication for PCV20 based on the 2023 ACIP guidelines (Supplemental Table 2).^16^ The index date was defined as the date of hospital admission or, if a respiratory specimen was collected on the day prior to admission, the date of that specimen collection, to capture cultures obtained in the emergency department before hospital admission. We excluded patients who received PCV20 within 30 days before the index date, as adequate immunity may not yet have developed. Patients were also excluded if they had received PPSV23 or PCV13 within the prior year, both PCV13 and PPSV23 within the prior five years (considered complete pneumococcal vaccination), or PCV15 or PCV21 at any time before the index date (Figure 1).^14,16^ Patients could contribute multiple pneumonia hospitalizations, provided each subsequent hospitalization event occurred at least 30 days after the prior discharge.²

**Figure 1:**
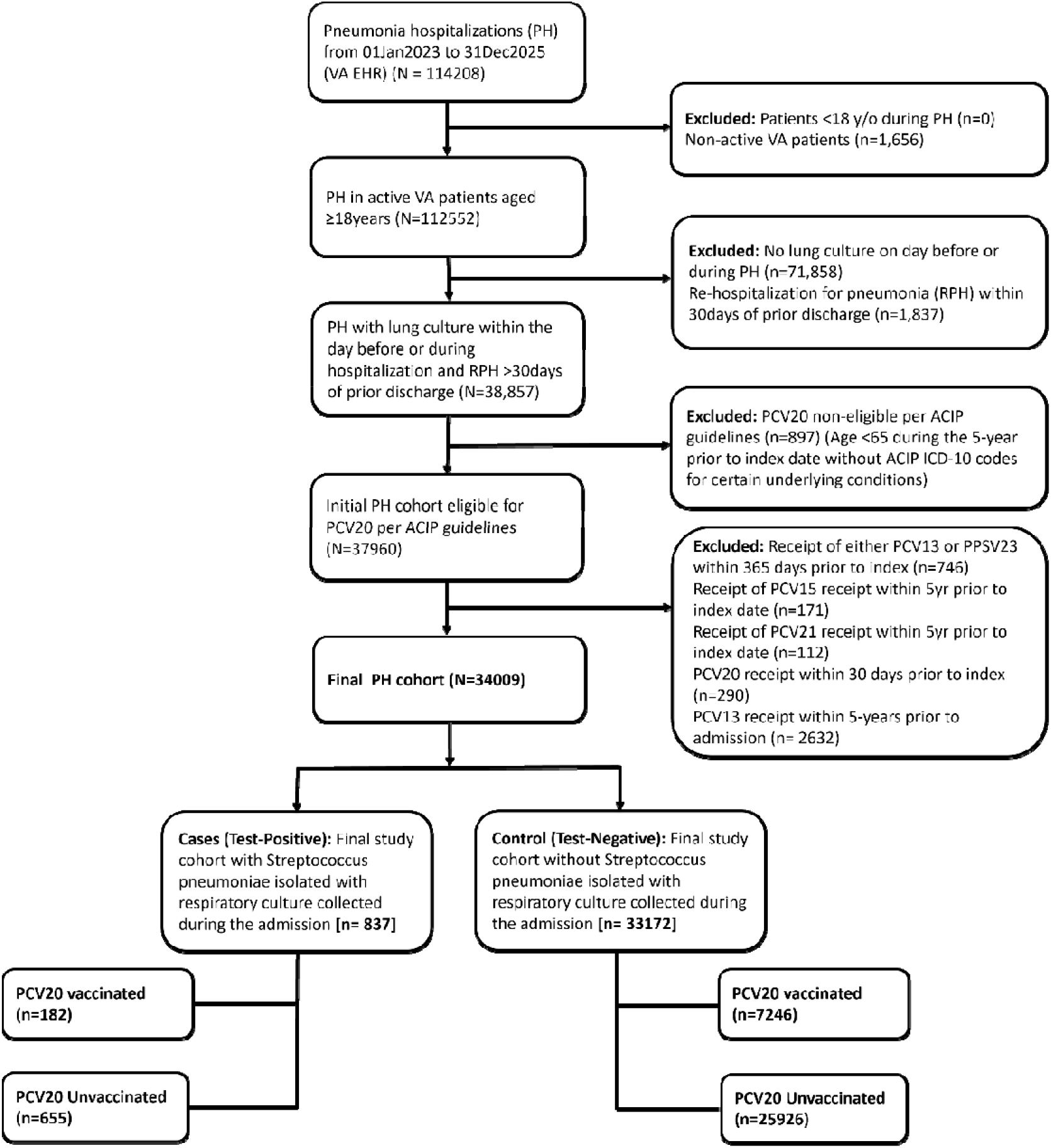
Study Population Flowchart

### Outcomes

Test-positive cases included patients hospitalized with pneumonia with a positive respiratory culture for *Streptococcus pneumoniae,* and test-negative controls whose respiratory cultures did not identify *Streptococcus pneumoniae*.

### Exposure

Patients who received PCV20 at least 30 days prior to hospital admission were classified as vaccinated,^2,12^ whereas those without any record of PCV20 receipt were classified as unvaccinated.

### Statistical Analyses

Odds ratios were estimated by comparing the odds of PCV20 vaccine receipt in test-positive cases and test-negative controls using a multivariable logistic regression model. VE was calculated as (1 - adjusted odds ratio) × 100. Covariate selection was informed by both clinical relevance and statistical criteria. Potential covariates with a p-value ≤0.25 in univariate analyses were included in the initial multivariable model. An automatic forward selection procedure was then applied, with statistically selected variables retained in the final model if they remained significant at p<0.05. Variables identified a priori as associated with pneumococcal disease risk or vaccination likelihood were included in all models, and additional variables that differed significantly between cases and controls were included to further control for confounding.

Variables considered for adjustment included age groups (18-64, 65-74, 75-84, ≥85 years), sex (male or female), race (Black, White, or other race), ethnicity (Hispanic or non-Hispanic), body mass index category (underweight, normal, overweight, obese), Charlson Comorbidity Index (0, 1, 2, 3, or ≥4), U.S. Census region (Northeast, Midwest, South, or West), chronic renal failure, malignancy, immunodeficiency, iatrogenic immunosuppression, leukemia, lymphoma, solid organ transplant, chronic heart disease, chronic lung disease, chronic liver disease, diabetes, smoking, alcoholism, low-risk status per CDC ACIP-defined risk criteria for PCV20 use in adults,^17^ prior PPSV23, influenza and COVID-19 vaccination in the 365 days prior to hospitalization, prior hospital admission, nursing home admission, intensive care unit admission, emergency department visit, number of outpatient visits [0-12, 13-24, or ≥25]), index year (2023-2025), and area deprivation index (least deprivation, moderate deprivation, high deprivation, or most deprivation/unknown) as a proxy for socioeconomic status. Other immunocompromising conditions were captured by a composite variable that included cochlear, asplenia, cerebrospinal fluid (CSF) leak, HIV infection, Hodgkin, multiple myeloma, nephrotic syndrome, sickle cell disease and other hemoglobinopathies. All logistic regression models were assessed for collinearity (variance inflation and tolerance) and model fit (Hosmer-Lemeshow).

VE was also estimated using inverse probability of treatment weighting (IPTW) to mitigate confounding by balancing baseline characteristics between vaccinated and unvaccinated individuals. Following a previously described approach for test-negative vaccine effectiveness studies,^18,19^ propensity score model for vaccination was fit (i.e., coefficients estimated) using logistic regression among test-negative controls only, and the resulting coefficients were subsequently used to generate predicted propensity scores for both test-positive cases and test-negative controls). Vaccinated patients were assigned stabilized weights equal to the marginal probability of vaccination divided by their estimated propensity score (P[A]/PS), while unvaccinated patients were weighted by the marginal probability of non-vaccination divided by the complement of their propensity score (P[1−A]/[1−PS]). Weights were winsorized (extreme weight trimming method based on percentile cutoff) at the 99.9th percentile to reduce the influence of extreme values.^19,20^ Covariate balance was assessed using standardized mean differences (SMD), with values <0.1 considered acceptable, and unbalanced covariates after weighting (SMD ≥0.1) were additionally adjusted for in the final weighted outcome model.

Variables included in each model can be found in Supplemental Tables 3a and 3b. All analyses were conducted using SAS version 9.4 (SAS Institute, Cary, North Carolina, USA).

### Subgroup Analyses

PCV20 VE was estimated by age group (18-49, 50-64, 65-74, 75-84, ≥85 years) and among those 65 years and older. Further, in those 65 years and older VE was assessed by CDC ACIP PCV20 risk-based recommendations,^17^ categorized as high-risk (patient with immunocompromised conditions), medium-risk (patients who were not immunocompromised but other chronic medical conditions), and low-risk (patients with neither immunocompromised or chronic medical conditions), and by U.S. census region (Northeast, Midwest, South, and West). The variables included in the high risk and medium risk categorizations are defined in Supplementary Table 2. Regional subgroup analyses were motivated by CDC surveillance data indicating a high and geographically concentrated burden of serotype 4 invasive pneumococcal disease, particularly in the Western region of the U.S.^8^

### Sensitivity Analyses

Consistent with recommendations from ACIP indicating that adults aged ≥65 years remain eligible for PCV20 regardless of prior PCV13 and PPSV23 receipt, we shortened the exclusion window for prior PCV13 receipt from 5 years in the primary analysis to 1 year in the sensitivity analysis.^21^ To assess the potential for unmeasured confounding in the IPTW-adjusted model, we conducted a negative control analysis using fracture and cataract as outcomes, defined from ICD-10-CM codes.

### Ethics Approval

This study complied with all relevant ethical regulations and was determined to be exempt by the VA Providence Healthcare System Institutional Review Board (IRB), with approval from the VAPHS Research and Development Committee. As a retrospective study of existing health records, informed consent was not required.

## Results

Among 34,009 individuals included in the analysis, 837 (2.5%) were test-positive cases and 33,172 (97.5%) were test-negative controls. Of the total study population, 21.8% (7,428/34,009) received PCV20 prior to the index date. The population was predominantly older (mean age 74.0 years [standard deviation (SD) 10.4]; median 75 years, [interquartile range (IQR): 69-80]; Table 1), male (95.9%), and White (74.4%), with 18.0% identifying as Black or African American.

**Table 1:** Baseline characteristics by case status, PCV20 vaccination status, unweighted and IPTW-weighted (N=34,009)

| Patient characteristics |  |  |  | Unweighted |  |  | IPTW Weighted Pseudo-Population |  |  |
| --- | --- | --- | --- | --- | --- | --- | --- | --- | --- |
|  | Overall | Cases (%) | Control (%) | PCV20 Unvaccinated N (%) | PCV20 Vaccinated N (%) | SMD | PCV20 Unvaccinated | PCV20 Vaccinated | SMD |
| **N** | 34009 | 837 (2.5) | 33172 (97.5) | 26581 (78.2) | 7428 (21.8) |  | 27928 (82.0) | 6127 (18.0) |  |
| **Age, years** |  |  |  |  |  | 0.421 |  |  | 0.006 |
| mean (SD) | 74.0 (10.4) | 72.7 (9.8) | 74.0 (10.5) | 74.8 (10.3) | 71.0 (10.5) |  | 74.1 (10.8) | 74.0 (9.2) |  |
| Median (IQR) | 75.0 (69.0, 80.0) | 74.0 (67.0-78.0) | 75.0 (69.0-80.0) | 76.0 (71.0-80.0) | 70.0 (65.0-78.0) |  | 75.0 (70.0-80.0) | 76.0 (68.0-80.0) |  |
| Minimum, maximum | 20, 106 | 25, 99 | 20, 106 | 22, 106 | 20, 103 |  | 22, 106 | 20, 103 |  |
| **Age groups (years)** |  |  |  |  |  | 0.367 |  |  | 0.024 |
| 18-49 | 946 | 20 (2.4) | 926 (2.8) | 730 (2.8) | 216 (2.9) |  | 777 (2.8) | 158 (2.6) |  |
| 50-64 | 4203 | 124 (14.8) | 4079 (12.3) | 2676 (10.1) | 1527 (20.6) |  | 3397 (12.2) | 742 (12.1) |  |
| 65-74 | 10140 | 283 (33.8) | 9857 (29.7) | 7319 (27.5) | 2821 (38.0) |  | 8324 (29.8) | 1776 (29.0) |  |
| 75-84 | 14576 | 340 (40.6) | 14236 (42.9) | 12365 (46.5) | 2211 (29.8) |  | 12016 (43.0) | 2708 (44.2) |  |
| ≥ 85 | 4144 | 70 (8.4) | 4074 (12.3) | 3491 (13.1) | 653 (8.8) |  | 3416 (12.2) | 744 (12.1) |  |
| **Sex** |  |  |  |  |  | 0.099 |  |  | 0.007 |
| Female | 1392 | 34 (4.1) | 1358 (4.1) | 967 (3.6) | 425 (5.7) |  | 1134 (4.1) | 254 (4.1) |  |
| Male | 32617 | 803 (95.9) | 31814 (95.9) | 25614 (96.4) | 7003 (94.3) |  | 26794 (95.9) | 5874 (95.9) |  |
| **Race** |  |  |  |  |  | 0.082 |  |  | 0.000 |
| Black or African American | 6133 | 117 (13.9) | 6016 (18.1) | 4687 (17.6) | 1446 (19.5) |  | 5035 (18.0) | 1088 (17.8) |  |
| White | 25311 | 661 (79.0) | 24650 (74.3) | 19976 (75.2) | 5335 (71.8) |  | 20776 (74.4) | 4578 (74.7) |  |
| American Indian, Alaska Native, Native Hawaiian, Pacific Islander, Asian, Unknown | 2565 | 59 (7.1) | 2506 (7.6) | 1918 (7.2) | 647 (8.7) |  | 2116 (7.6) | 461 (7.5) |  |
| **Ethnicity** |  |  |  |  |  | 0.015 |  |  | 0.007 |
| Hispanic/Latino | 2233 | 41 (4.9) | 2192 (6.6) | 1724 (6.5) | 509 (6.9) |  | 1845 (6.6) | 405 (6.6) |  |
| *Non-Hispanic/Latino/Unknown* | 31776 | 796 (95.1) | 30980 (93.4) | 24857 (93.5) | 6919 (93.2) |  | 26083 (93.4) | 5723 (93.4) |  |
| **U.S. geographic region** |  |  |  |  |  | 0.052 |  |  | 0.031 |
| *West* | 6921 | 174 (20.8) | 6747 (20.3) | 5353 (20.1) | 1568 (21.1) |  | 5696 (20.4) | 1266 (20.7) |  |
| *Midwest* | 7044 | 252 (30.1) | 6792 (20.5) | 5618 (21.1) | 1426 (19.2) |  | 5782 (20.7) | 1246 (20.3) |  |
| *Northeast* | 5013 | 162 (19.4) | 4851 (14.6) | 3932 (14.8) | 1081 (14.6) |  | 4115 (14.7) | 900 (14.7) |  |
| *South* | 15031 | 249 (29.8) | 14782 (44.6) | 11678 (43.9) | 3353 (45.1) |  | 12336 (44.2) | 2716 (44.3) |  |
| **Risk groups and conditions** |  |  |  |  |  | 0.095 |  |  | 0.020 |
| **High risk (IC)** | 15011 | 302 (36.1) | 14709 (44.3) | 11893 (44.7) | 3118 (42.0) |  | 12354 (44.2) | 2748 (44.8) |  |
| *Chronic renal failure* | 2884 | 33 (3.9) | 2851 (8.6) | 2371 (8.9) | 513 (6.9) | 0.075 | 2375 (8.5) | 572 (9.3) | 0.029 |
| *Malignancy* | 11766 | 248 (29.6) | 11518 (34.7) | 9378 (35.3) | 2388 (32.2) | 0.066 | 9655 (34.6) | 2131 (34.8) | 0.004 |
| *Immunodeficiency* | 868 | 17 (2.0) | 851 (2.6) | 663 (2.5) | 205 (2.8) | 0.017 | 709 (2.5) | 158 (2.6) | 0.002 |
| *Iatrogenic immunosuppression* | 1395 | 32 (3.8) | 1363 (4.1) | 1054 (4.0) | 341 (4.6) | 0.031 | 1144 (4.1) | 261 (4.3) | 0.008 |
| *Leukemia* | 915 | 14 (1.7) | 901 (2.7) | 743 (2.8) | 172 (2.3) | 0.030 | 757 (2.7) | 178 (2.9) | 0.012 |
| *Lymphoma* | 854 | 15 (1.8) | 839 (2.5) | 692 (2.6) | 162 (2.2) | 0.028 | 705 (2.5) | 166 (2.7) | 0.011 |
| *Solid organ transplant* | 695 | 9 (1.1) | 686 (2.1) | 507 (1.9) | 188 (2.5) | 0.042 | 562 (2.0) | 118 (1.9) | 0.006 |
| *Other IC conditions<sup>®</sup>* | 1381 | 27 (3.2) | 1354 (4.1) | 1038 (3.9) | 343 (4.6) | 0.035 | 1137 (4.1) | 276 (4.5) | 0.021 |
| **Medium risk (CMC)** | 18029 | 511 (61.1) | 17518 (52.8) | 13872 (52.2) | 4157 (56.0) |  | 14781 (52.9) | 3206 (52.3) |  |
| *Alcohol Use* | 8452 | 219 (26.2) | 8233 (24.8) | 6318 (23.8) | 2134 (28.7) | 0.113 | 6923 (24.8) | 1525 (24.9) | 0.002 |
| *Smoking* | 26124 | 714 (85.3) | 25410 (76.6) | 20319 (76.4) | 5805 (78.2) | 0.041 | 21445 (76.8) | 4686 (76.5) | 0.007 |
| *Diabetes* | 15047 | 316 (36.5) | 14731 (44.4) | 11726 (44.1) | 3321 (44.7) | 0.012 | 12360 (44.3) | 2677 (43.7) | 0.011 |
| *Chronic heart disease* | 16670 | 342 (40.9) | 16328 (49.2) | 13121 (49.4) | 3549 (47.8) | 0.032 | 13696 (49.0) | 3065 (50.0) | 0.020 |
| *Chronic lung disease* | 20141 | 546 (65.2) | 19595 (59.1) | 15468 (58.2) | 4673 (62.9) | 0.097 | 16539 (59.2) | 3638 (59.4) | 0.003 |
| *Chronic liver disease* | 2286 | 55 (6.6) | 2231 (6.7) | 1675 (6.3) | 611 (8.2) | 0.074 | 1863 (6.7) | 402 (6.6) | 0.005 |
| **Low risk** | 969 | 24 (2.9) | 945 (2.9) | 816 (3.1) | 153 (2.1) |  | 793 (2.8) | 174 (2.8) |  |
| **Body mass index** |  |  |  |  |  | 0.069 |  |  | 0.030 |
| *Underweight (<18.5)* | 1368 | 44 (5.3) | 1324 (4.0) | 1076 (4.1) | 292 (3.9) |  | 1115 (4.0) | 240 (3.9) |  |
| *Normal (18.5–24.9)\*\** | 16225 | 461 (55.1) | 15764 (47.5) | 12889 (48.5) | 3336 (44.9) |  | 13391 (48.0) | 2951 (48.2) |  |
| *Overweight (25.0–29.9)* | 6091 | 148 (17.7) | 5943 (17.9) | 4789 (18.0) | 1302 (17.5) |  | 5020 (18.0) | 1061 (17.3) |  |
| *Obese (≥30.0)* | 10325 | 184 (22.0) | 10141 (30.6) | 7827 (29.5) | 2498 (33.6) |  | 8402 (30.1) | 1877 (30.6) |  |
| **Healthcare Utilization, 1 year** |  |  |  |  |  |  |  |  |  |
| *Hospitalization* | 17901 | 369 (44.1) | 17532 (52.9) | 13787 (51.9) | 4114 (55.4) | 0.071 | 14713 (52.7) | 3274 (53.4) | 0.015 |
| *Nursing home admission* | 1855 | 38 (4.5) | 1817 (5.5) | 1321 (5.0) | 534 (7.2) | 0.093 | 1532 (5.5) | 355 (5.8) | 0.014 |
| *Intensive care unit admission* | 5737 | 83 (9.9) | 5654 (17.0) | 4309 (16.2) | 1428 (19.2) | 0.079 | 4719 (16.9) | 1072 (17.5) | 0.016 |
| *Emergency department visit* | 25756 | 593 (70.9) | 25163 (75.9) | 20027 (75.3) | 5729 (77.1) | 0.042 | 21132 (75.7) | 4660 (76.1) | 0.009 |
| *Number of outpatient visits* |  |  |  |  |  | 0.141 |  |  | <0.001 |
| *0-12* | 4858 | 153 (18.3) | 4705 (14.2) | 4072 (15.3) | 786 (10.6) |  | 4000 (14.3) | 855 (14.0) |  |
| *13-24* | 6410 | 196 (23.4) | 6214 (18.7) | 5160 (19.4) | 1250 (16.8) |  | 5261 (18.8) | 1171 (19.1) |  |
| *25+* | 22741 | 488 (58.3) | 22253 (67.1) | 17349 (65.3) | 5392 (72.6) |  | 18668 (66.8) | 4100 (66.9) |  |
| **Index Year** |  |  |  |  |  | 0.373 |  |  | 0.024 |
| 2023 | 9894 | 239 (28.6) | 9655 (29.1) | 8545 (32.2) | 1349 (18.2) |  | 8122 (29.1) | 1761 (28.7) |  |
| 2024 | 11719 | 257 (30.7) | 11462 (34.6) | 9204 (34.6) | 2515 (33.9) |  | 9625 (34.5) | 2125 (34.7) |  |
| 2025 | 12396 | 341 (40.7) | 12055 (36.3) | 8832 (33.2) | 3564 (48.0) |  | 10181 (36.5) | 2242 (36.6) |  |
| **Prior vaccine receipt (before index)** |  |  |  |  |  |  |  |  |  |
| *PPSV23, prior 5 years* | 6703 | 151 (18.0) | 6552 (19.8) | 5276 (19.9) | 1427 (19.2) | 0.016 | 5552 (19.9) | 1323 (21.6) | 0.042 |
| Influenza vaccination, prior 365 days | 20591 | 497 (59.4) | 20094 (60.6) | 15239 (57.3) | 5352 (72.1) | 0.312 | 16865 (60.4) | 3571 (58.3) | 0.043 |
| COVID-19 vaccination, prior 365 days | 12151 | 300 (35.8) | 11851 (35.7) | 8960 (33.7) | 3191 (43.0) | 0.191 | 9958 (35.7) | 2158 (35.2) | 0.009 |
| **Charlson comorbidity index** |  |  |  |  |  | 0.104 |  |  | <0.001 |
| 0 | 3483 | 93 (11.1) | 3390 (10.2) | 2854 (10.7) | 629 (8.5) |  | 2857 (10.2) | 612 (10.0) |  |
| 1 | 6321 | 212 (25.3) | 6109 (18.4) | 4895 (18.4) | 1426 (19.2) |  | 5199 (18.6) | 1148 (18.7) |  |
| 2 | 5459 | 146 (17.4) | 5313 (16.0) | 4182 (16.0) | 1277 (17.2) |  | 4472 (16.0) | 997 (16.3) |  |
| 3 | 4655 | 116 (13.9) | 4539 (13.7) | 3603 (13.6) | 1052 (14.2) |  | 3811 (13.7) | 829 (13.5) |  |
| 4+ | 14091 | 270 (32.3) | 13821 (41.7) | 11047 (41.6) | 3044 (41.0) |  | 11588 (41.5) | 2542 (41.5) |  |
| **Area Deprivation Index** |  |  |  |  |  | <0.001 |  |  | 0.027 |
| Least deprived | 7937 | 182 (21.7) | 7755 (23.4) | 6194 (23.3) | 1743 (23.5) |  | 6535 (23.4) | 1420 (23.2) |  |
| Moderate deprivation | 7833 | 207 (24.7) | 7626 (23.0) | 6103 (23.2) | 1730 (23.3) |  | 6446 (23.1) | 1430 (23.4) |  |
| High deprivation | 8079 | 228 (27.2) | 7851 (23.7) | 6311 (23.7) | 1768 (23.8) |  | 6596 (23.6) | 1440 (23.5) |  |
| Most deprivation/Unknown | 10160 | 220 (26.3) | 9940 (30.0) | 7973 (30.0) | 2187 (29.4) |  | 8351 (29.9) | 1837 (30.0) |  |

SD, standard deviation; IQR, interquartile range; SMD, standardized mean difference; IPTW, inverse probability of treatment weighting; IC: Immunocompromised Conditions; CMC: Chronic Medical Conditions PCV13, 13-valent pneumococcal conjugate vaccine; PCV20, 20-valent pneumococcal conjugate vaccine; PPSV23, pneumococcal polysaccharide vaccine; <sup>2</sup> Conditions with small cell count adjusted as a group variable (<2%; Cochlear, asplenia, cerebrospinal fluid (CSF) leak, HIV infection, Hodgkin, multiple myeloma, nephrotic syndrome, sickle cell disease and other hemoglobinopathies); \*\*Missing body mass index categorized as Normal (n=17).

Compared with test-negative controls, test-positive cases were slightly younger (mean age 72.7 vs 74.0 years) and had a lower prevalence of several comorbid conditions, including chronic renal failure (3.9% vs 8.6%), malignancy (29.6% vs 34.7%), diabetes (36.5% vs 44.4%), and chronic heart disease (40.9% vs 49.2%). Cases were also more likely to have chronic lung disease (65.2% vs 59.1%) and a history of smoking (85.3% vs 76.6%). Additional differences in demographic and clinical characteristics between cases and controls are presented in Table 1.

VE against pneumococcal pneumonia hospitalizations was demonstrated among adults aged ≥65 years (adjusted 20.4%; 95% CI: 2.1% to 35.3%). Within this age group, significant protection was observed among individuals with chronic medical conditions (medium risk group 24.4%; 95% CI: 0.7% to 42.4%). VE was highest in the Western U.S. (39.6%; 95% CI: 3.2% to 62.3%), whereas estimates in other regions were lower and not statistically significant.

Although the overall VE estimate was not statistically significant (8.8%; 95% CI: −8.8% to 23.6%), and confidence intervals for most age-, risk-, and region-specific estimates included the null, the direction of the point estimates was generally consistent with a protective association.

An exception was observed among adults aged 50-64 years, for whom VE estimates were negative (−63.1%; 95% CI: −141.1% to −10.3%).

Findings were generally consistent across multivariable and IPTW-adjusted analyses. In the IPTW-adjusted model, VE remained significant among individuals 65 years and older with chronic medical conditions (medium risk group 23.8%; 95% CI: 0.8% to 41.4%) and in the Western U.S. (49.6%; 95% CI: 16.5% to 69.6%) (Table 2).

**Table 2:** PCV20 effectiveness against pneumococcal pneumonia hospitalization among adults without prior PCV13 receipt (N=34,009)

|  |  | Unadjusted VE (95% CI) | Multivariable Model (95% CI) | IPTW adjusted VE (95% CI) |
| --- | --- | --- | --- | --- |
| **Age groups** | 18 and above (34009) | 0.58 (-17.41, 15.81) | 8.82 (-8.76, 23.55) | 8.39 (-9.99, 23.71) |
|  | 18-49 years (946) | 63.01 (-60.63, 91.48) | 41.20 (-187.43, 87.97) | 74.15 (-228.39, 97.97) |
|  | 50-64 years (4203) | **-56.14 (-123.46, -9.10)** | **-63.07 (-141.14, -10.28)** | **-71.54 (-166.55, -10.40)** |
|  | 65-74 years (10140) | 13.45 (-13.65, 34.08) | 10.57 (-17.73, 32.06) | 8.42 (-19.19, 29.63) |
|  | 75-84 years (14576) | 14.99 (-16.71, 38.09) | 18.12 (-13.29, 40.82) | 13.70 (-18.26, 37.02) |
|  | 85+ years (4144) | 50.34 (-15.15, 78.58) | 44.82 (-30.60, 76.68) | 47.54 (-19.76, 77.02) |
| **≥65 years** | 65 + years (28860) | 13.18 (-5.79, 28.75) | **20.38 (2.07, 35.27)** | 18.02 (-0.10, 32.86) |
|  | **Risk group** |  |  |  |
|  | High risk (IC) (13318) | 5.18 (-30.05, 30.86) | 13.82 (-19.86, 38.03) | 6.74 (-27.86, 31.98) |
|  | Medium risk (CMC) (14725) | 18.81 (-5.20, 37.35) | **24.36 (0.71, 42.37)** | **23.77 (0.77, 41.44)** |
|  | Low risk (817) | 15.37 (-190.29, 75.32) | 15.56 (-239.56, 79.00) | 42.28 (-142.24, 86.25) |
|  | **U.S. census regions** |  |  |  |
|  | West (5886) | 29.11 (-11.19, 54.80) | **39.57 (3.16, 62.29)** | **49.57 (16.46, 69.56)** |
|  | Northeast (4303) | 10.12 (-41.81, 43.03) | 12.68 (-40.92, 45.90) | 13.88 (-37.03, 45.87) |
|  | South (12618) | 2.94 (-38.14, 31.80) | 8.48 (-32.56, 36.82) | 2.16 (-38.67, 30.97) |
|  | Midwest (6053) | 7.81 (-32.14, 35.68) | 21.85 (-14.35, 46.59) | 15.57 (-21.71, 41.43) |

IPTW, inverse probability of treatment weighted; IC: Immunocompromised Conditions; CMC: Chronic Medical Conditions
Bold indicates statistically significant, >0 indicates vaccine effectiveness, <0 indicates vaccine ineffectiveness.

Results were also consistent in the sensitivity analysis that shortened the exclusion window for prior PCV13 receipt from 5 years to 1 year. Among adults aged ≥65 years, VE was 18.0% (95% CI: −0.2% to 32.9%), with similar patterns observed across age-, risk-, and region-specific estimates, including higher point estimates among individuals with chronic medical conditions and in the Western U.S. (Supplemental Table 4).

Negative control outcome analyses based on the IPTW-adjusted model demonstrated no meaningful association between PCV20 vaccination and fracture (VE −0.3%; 95% CI: −11.7% to 9.9%) or cataract (VE 3.2%; 95% CI: −3.1% to 9.2%). Estimates were similarly close to the null across age-, risk-, and region-specific estimates with no consistent pattern of association (Supplemental Table 5).

## Discussion

This study provides, to our knowledge, the first real-world estimate of PCV20 effectiveness against culture-confirmed pneumococcal pneumonia hospitalizations. In this test-negative analysis of VA adults, PCV20 was associated with a 20.4% reduction in the odds of pneumococcal pneumonia hospitalization among adults aged ≥65 years. Notably, vaccine effectiveness was substantially higher in the Western U.S. (39.6%), suggesting potential geographic heterogeneity in vaccine impact. These findings align with CDC Active Bacterial Core surveillance data demonstrating a disproportionate burden of serotype 4 disease in the Western U.S.^8^ As serotype 4 is included in PCV20,^22^ regional differences in circulating serotypes may partially explain observed differences in vaccine effectiveness.^8^ More broadly, our findings suggest that regional serotype epidemiology may influence the population-level impact of pneumococcal conjugate vaccines and highlight the importance of ongoing serotype surveillance when evaluating vaccine effectiveness and informing vaccination strategies.^8^

Our estimate of 20.4% VE against culture-confirmed pneumococcal pneumonia hospitalizations among adults aged ≥65 years was similar in magnitude to the 23.3% VE against all-cause pneumonia hospitalizations reported in a recent Medicare cohort study .^11^ The Medicare study identified pneumococcal pneumonia using ICD-10-CM codes without microbiologic confirmation. The authors noted that the incidence of pneumococcal pneumonia identified from diagnosis codes was similar to the incidence of invasive pneumococcal disease identified from diagnosis codes, and substantially lower than previously reported population-based estimates of pneumococcal pneumonia. They suggested that diagnosis code-based definitions of pneumococcal pneumonia may preferentially capture severe or bacteremic cases, for which clinical confirmation and documentation of a specific pneumococcal etiology are more likely to be pursued.^11^ Consequently, concerns regarding the validity and completeness of diagnosis code-defined pneumococcal pneumonia led the authors to focus their primary effectiveness analyses on all-cause pneumonia hospitalizations rather than pneumococcal pneumonia specifically. By using culture-confirmed pneumococcal pneumonia, our study was able to directly estimate PCV20 effectiveness against microbiologically confirmed pneumococcal disease while reducing outcome misclassification.

The negative VE estimates observed among adults aged 50-64 years should be interpreted cautiously. During most of the study period, pneumococcal vaccination in this age group was recommended only for individuals with underlying risk conditions.^14,16^ However, vaccine uptake among eligible at-risk adults remained relatively low, resulting in a vaccinated population that likely represented a particularly high risk subset of patients with greater underlying susceptibility to pneumococcal disease. Residual confounding by indication may therefore explain the observed negative estimates despite adjustment for measured risk factors. In October 2024, ACIP expanded recommendations to all adults aged ≥50 years, in part to improve vaccine coverage among at-risk adults.^23^ Future studies conducted under this age-based recommendation may provide a less confounded assessment of PCV20 effectiveness in adults aged 50-64 years.

The test-negative design reduces confounding by healthcare-seeking behavior because both cases and controls sought care and underwent respiratory culture testing.^24,25^ In addition, IPTW achieved good balance across measured covariates, including index year, reducing the potential for temporal confounding. This contrasts with the recent Medicare cohort study, in which substantial imbalance in index year remained after weighting.^11^ Negative control outcome analyses in our study yielded estimates close to the null for fracture and cataract, whereas the Medicare study reported significant associations for at least one negative control outcome.^11^ Collectively, these findings suggest that the observed VE estimates are unlikely to be explained by major sources of bias.

Our study is subject to several limitations. Given the observational nature of the study, the findings should be interpreted as estimates of association and warrant confirmation in future studies. While the test-negative design is well-suited for evaluating vaccine effectiveness by reducing bias related to healthcare-seeking and testing behavior, it remains susceptible to selection bias.^26,27^ Residual confounding by unmeasured factors cannot be excluded despite adjustment for measured clinical and sociodemographic characteristics and the use of inverse probability of treatment weighting. Differences in health-seeking behavior, access to care, and adherence to preventive measures may influence both vaccination status and the likelihood of pneumococcal pneumonia hospitalization. While VA vaccination records are supplemented with information from state immunization registries and patient reports, incomplete capture of vaccinations received outside the VA could have resulted in some misclassification of vaccination status. Such misclassification would likely bias estimates toward the null. Restricting outcomes to culture-confirmed pneumococcal pneumonia improved diagnostic specificity but may have reduced representativeness by limiting inclusion to patients who underwent microbiologic testing. Because testing is often pursued in patients with more severe illness, the study population may overrepresent severe pneumococcal pneumonia. We considered this tradeoff acceptable given the potential for substantial outcome misclassification when pneumococcal pneumonia is identified using administrative diagnosis codes alone. The absence of serotype data precludes estimation of serotype-specific vaccine effectiveness and limits inference regarding the contribution of serotype distribution, including serotype 4, to observed regional differences. Finally, generalizability may be limited. The VA population is predominantly older, male, and medically complex, and patterns of healthcare utilization may differ from patients in other healthcare systems.

In conclusion, PCV20 provided real-world protection against culture-confirmed pneumococcal pneumonia hospitalization among adults aged ≥65 years, representing, to our knowledge, the first direct evidence of PCV20 effectiveness against microbiologically confirmed pneumococcal pneumonia. The higher effectiveness observed in regions with a greater burden of serotype 4 disease suggests that local serotype epidemiology may be an important determinant of vaccine impact. Future studies incorporating serotype-specific outcomes and evaluating effectiveness among adults aged 50-64 years under the updated age-based vaccination recommendations will be important to further define the population-level impact of PCV20.

## Data Availability

Identifiable protected health data from the Veterans Health Administration support the findings of this study and cannot be made publicly available. Privacy regulations do not allow for open sharing of the individual-level data used in this study. After verifying de-identification, the Veterans Health Administration may approve sharing some of the study data, but it may not include all final study data. Requests for such data should be made to the corresponding author and are subject to approvals by the ethics board, privacy office, and the information systems and security office

## Acknowledgement

**Potential Conflict of Interest:** AO, SK, and SC have no conflict of interest to declare. ARC has received research funding from Pfizer and Bayer for research unrelated to this study.

**Availability of Data and Materials:** Identifiable protected health data from the Veterans Health Administration support the findings of this study and cannot be made publicly available. Privacy regulations do not allow for open sharing of the individual-level data used in this study. After verifying de-identification, the Veterans Health Administration may approve sharing some of the study data, but it may not include all final study data. Requests for such data should be made to the corresponding author and are subject to approvals by the ethics board, privacy office, and the information systems and security office.

**Funding/Support:** No external funding was received for this study.

**Author Contributions:** Conceptualization: AO, ARC, SK, SC; IRB submission, study design, and data acquisition: AO, ARC, VVL; Data analysis: AO, VVL; Drafting of the manuscript: AO, ARC, SK, SC; Critical revision of the manuscript for important intellectual content: AO, ARC, SC, SK.

## References

1. CDC. Pneumococcal Disease Surveillance and Trends. Pneumococcal Disease. January 21, 2025. Accessed April 1, 2025. https://www.cdc.gov/pneumococcal/php/surveillance/index.html

2. McLaughlin JM, Jiang Q, Isturiz RE, et al. Effectiveness of 13-Valent Pneumococcal Conjugate Vaccine Against Hospitalization for Community-Acquired Pneumonia in Older US Adults: A Test-Negative Design. Clin Infect Dis. 2018;67(10):1498–1506. doi:10.1093/cid/ciy312

3. Heo JY, Seo YB, Choi WS, et al. Effectiveness of Pneumococcal Vaccination Against Pneumococcal Pneumonia Hospitalization in Older Adults: A Prospective, Test-Negative Study. J Infect Dis. 2022;225(5):836–845. doi:10.1093/infdis/jiab474

4. Dion CF, Ashurst JV. Streptococcus pneumoniae. In: StatPearls. StatPearls Publishing; 2025. Accessed April 1, 2025. http://www.ncbi.nlm.nih.gov/books/NBK470537/

5. CDC. Chapter 17: Pneumococcal Disease. Epidemiology and Prevention of Vaccine-Preventable Diseases. December 9, 2024. Accessed March 23, 2025. https://www.cdc.gov/pinkbook/hcp/table-of-contents/chapter-17-pneumococcal-disease.html

6. Regunath H, Oba Y. Community-Acquired Pneumonia. In: StatPearls. StatPearls Publishing; 2026. Accessed June 29, 2026. http://www.ncbi.nlm.nih.gov/books/NBK430749/

7. Grant LR, Slack MPE, Theilacker C, et al. Distribution of Serotypes Causing Invasive Pneumococcal Disease in Children From High-Income Countries and the Impact of Pediatric Pneumococcal Vaccination. Clin Infect Dis. 2022;76(3):e1062–e1070. doi:10.1093/cid/ciac475

8. CDC. Summary of Risk-based Pneumococcal Vaccination Recommendations. Pneumococcal Disease. February 25, 2026. Accessed April 2, 2026. https://www.cdc.gov/pneumococcal/hcp/vaccine-recommendations/risk-indications.html

9. About Pneumococcal Vaccines: For Providers | CDC. September 12, 2024. Accessed March 23, 2025. https://www.cdc.gov/vaccines/vpd/pneumo/hcp/about-vaccine.html

10. Essink B, Sabharwal C, Cannon K, et al. Pivotal Phase 3 Randomized Clinical Trial of the Safety, Tolerability, and Immunogenicity of 20-Valent Pneumococcal Conjugate Vaccine in Adults Aged ≥18 Years. Clin Infect Dis. 2022;75(3):390-398. doi:10.1093/cid/ciab990

11. Miles AC, Vojicic J, Peyrani P, et al. Real-world effectiveness of 20-valent pneumococcal conjugate vaccine against all-cause outcomes among Medicare beneficiaries aged 65 years and older in the USA: a retrospective cohort study. The Lancet Infectious Diseases. 2026;0(0). doi:10.1016/S1473-3099(26)00115-5

12. Le D, Chang A, Grams ME, Coresh J, Ishigami J. Pneumococcal vaccination effectiveness (PCV13 and PPSV23) in individuals with and without reduced kidney function: a test-negative design study. Clinical Kidney Journal. 2024;17(6):sfae145. doi:10.1093/ckj/sfae145

13. Corporate Data Warehouse (CDW). January 11, 2023. Accessed April 2, 2026. https://www.hsrd.research.va.gov/for_researchers/cdw.cfm

14. Kobayashi M. Use of 15-Valent Pneumococcal Conjugate Vaccine and 20-Valent Pneumococcal Conjugate Vaccine Among U.S. Adults: Updated Recommendations of the Advisory Committee on Immunization Practices — United States, 2022. MMWR Morb Mortal Wkly Rep. 2022;71. doi:10.15585/mmwr.mm7104a1

15. Clinical Classifications Software Refined (CCSR) for ICD-10-CM Diagnoses. Accessed May 7, 2025. https://hcup-us.ahrq.gov/toolssoftware/ccsr/dxccsr.jsp#download

16. Kobayashi M. Pneumococcal Vaccine for Adults Aged ≥19 Years: Recommendations of the Advisory Committee on Immunization Practices, United States, 2023. MMWR Recomm Rep. 2023;72. doi:10.15585/mmwr.rr7203a1

17. Kobayashi M. Expanded Recommendations for Use of Pneumococcal Conjugate Vaccines Among Adults Aged ≥50 Years: Recommendations of the Advisory Committee on Immunization Practices — United States, 2024. MMWR Morb Mortal Wkly Rep. 2025;74. doi:10.15585/mmwr.mm7401a1

18. Månsson R, Joffe MM, Sun W, Hennessy S. On the Estimation and Use of Propensity Scores in Case-Control and Case-Cohort Studies. Am J Epidemiol. 2007;166(3):332–339. doi:10.1093/aje/kwm069

19. Thompson MG, Stenehjem E, Grannis S, et al. Effectiveness of Covid-19 Vaccines in Ambulatory and Inpatient Care Settings. N Engl J Med. 2021;385(15):1355–1371. doi:10.1056/NEJMoa2110362

20. Lee BK, Lessler J, Stuart EA. Weight Trimming and Propensity Score Weighting. PLOS ONE. 2011;6(3):e18174. doi:10.1371/journal.pone.0018174

21. CDC. Pneumococcal Vaccine Recommendations. Pneumococcal Disease. February 25, 2026. Accessed April 15, 2026. https://www.cdc.gov/pneumococcal/hcp/vaccine-recommendations/index.html

22. CDC. Pneumococcal Vaccination: Information for Health Care Providers. Vaccines & Immunizations. January 23, 2026. Accessed July 2, 2026. https://www.cdc.gov/vaccines/hcp/by-disease/pneumo.html

23. Kobayashi M. Expanded Recommendations for Use of Pneumococcal Conjugate Vaccines Among Adults Aged ≥50 Years: Recommendations of the Advisory Committee on Immunization Practices — United States, 2024. MMWR Morb Mortal Wkly Rep. 2025;74. doi:10.15585/mmwr.mm7401a1

24. Li G, Gerlovin H, Figueroa Muñiz MJ, et al. Comparison of the test-negative design and cohort design with explicit target trial emulation for evaluating Covid-19 vaccine effectiveness. Epidemiology. 2024;35(2):137–149. doi:10.1097/EDE.0000000000001709

25. Dean NE, Hogan JW, Schnitzer ME. Covid-19 Vaccine Effectiveness and the Test-Negative Design. New England Journal of Medicine. 2021;385(15):1431–1433. doi:10.1056/NEJMe2113151

26. De Serres G, Skowronski DM, Wu XW, Ambrose CS. The test-negative design: validity, accuracy and precision of vaccine efficacy estimates compared to the gold standard of randomised placebo-controlled clinical trials. Euro Surveill. 2013;18(37):20585. doi:10.2807/1560-7917.es2013.18.37.20585

27. Jackson ML, Phillips CH, Benoit J, et al. The impact of selection bias on vaccine effectiveness estimates from test-negative studies. Vaccine. 2018;36(5):751–757. doi:10.1016/j.vaccine.2017.12.022

