## Supplemental File for "Effectiveness of 20-valent pneumococcal conjugate vaccine against pneumococcal pneumonia hospitalizations among U.S. adults: Test-Negative Design"

**Supplemental Material**

Supplemental Table 1: Pneumonia International Classification of Diseases (ICD), 10^th^ Edition, Diagnosis Codes^1^

| **ICD-10 Code** | **ICD-10 Code Description** |
| --- | --- |
| J1000 | Influenza due to identified seasonal influenza virus with pneumonia |
| J1001 | Influenza due to other identified influenza virus with pneumonia |
| J1008 | Influenza with other respiratory manifestations |
| J1100 | Influenza due to unidentified influenza virus with pneumonia |
| J1108 | Influenza with other respiratory manifestations, virus not identified |
| J120 | Adenoviral pneumonia |
| J121 | Respiratory syncytial virus pneumonia |
| J122 | Parainfluenza virus pneumonia |
| J123 | Human metapneumovirus pneumonia |
| J1281 | Pneumonia due to SARS-associated coronavirus |
| J1289 | Other viral pneumonia |
| J129 | Viral pneumonia, unspecified |
| J13 | Pneumonia due to Streptococcus pneumoniae |
| J14 | Pneumonia due to Haemophilus influenzae |
| J150 | Pneumonia due to Klebsiella pneumoniae |
| J151 | Pneumonia due to Pseudomonas |
| J1520 | Pneumonia due to staphylococcus, unspecified |
| J15211 | Pneumonia due to methicillin susceptible Staphylococcus aureus |
| J15212 | Pneumonia due to methicillin resistant Staphylococcus aureus |
| J1529 | Pneumonia due to other staphylococcus |
| J153 | Pneumonia due to streptococcus, group B |
| J154 | Pneumonia due to other streptococci |
| J155 | Pneumonia due to Escherichia coli |
| J156 | Pneumonia due to other Gram-negative bacteria |
| J157 | Pneumonia due to Mycoplasma pneumoniae |
| J158 | Pneumonia due to other specified bacteria |
| J159 | Unspecified bacterial pneumonia |
| J160 | Chlamydial pneumonia |
| J168 | Pneumonia due to other specified infectious organisms |
| J17 | Pneumonia in diseases classified elsewhere |
| J180 | Bronchopneumonia, unspecified organism |
| J181 | Lobar pneumonia, unspecified organism |
| J188 | Other pneumonia, unspecified organism |
| J189 | Pneumonia, unspecified organism |
| A221 | Pulmonary anthrax |
| A3701 | Whooping cough due to Bordetella pertussis with pneumonia |
| A3711 | Whooping cough due to Bordetella parapertussis with pneumonia |
| A3781 | Whooping cough due to other Bordetella species with pneumonia |
| A3791 | Whooping cough, unspecified species with pneumonia |
| A481 | Legionnaires’ disease |
| B250 | Cytomegaloviral pneumonitis |
| B440 | Invasive pulmonary aspergillosis |
| B7781 | Ascariasis pneumonia |

Supplemental Table 2: PCV20 Risk-Based Conditions in Adults^2^

| **Risk Factors/Medical Conditions** | **ICD-10 Codes or Health Factors** |
| --- | --- |
| **High risk (Immunocompromised Conditions)** | |
| Chronic renal failure | N18.4-N18.6 (Chronic kidney disease stage 4-) |
| Nephrotic syndrome | N04.x (Nephrotic syndrome) |
| Immunodeficiency | D80- D84 (Immunodeficiency disorders) |
| Iatrogenic immunosuppression | Z79.52 (Long-term use of immunosuppressants), medication records (e.g., corticosteroids, methotrexate) |
| Generalized malignancy | C80.0 (Disseminated malignancy), C00-C97 (Malignant neoplasms) |
| HIV infection | B20 - B24 (HIV disease) |
| Hodgkin disease | C81.x (Hodgkin lymphoma) |
| Leukemia | C91-C95 (Leukemias) |
| Lymphoma | C81-C85 (Lymphomas) |
| Multiple myeloma | C90.0 (Multiple myeloma) |
| Solid organ transplants | Z94.x (Transplanted organ and tissue status) |
| Congenital or acquired asplenia | D73.0 (Asplenia), D73.89 (Other splenic diseases), Z90.81 (Acquired absence of spleen) |
| Sickle cell disease or other hemoglobinopathies | D57.x (Sickle cell disorders, except D57.3), D58.x (Other hereditary hemolytic anemias) |
| Cochlear implant | Z96.21 (Presence of cochlear implant) |
| CSF leak | G96.0 (Cerebrospinal fluid leak) |
| **Medium risk (Chronic Medical Conditions)** | |
| Alcoholism | F10.1x (Alcohol abuse), F10.2x (Alcohol dependence), Alcohol Use = ‘Yes’ (VA Health Factor) |
| Cigarette smoking | F17.2x (Nicotine dependence), Z72.0 (Tobacco use), Smoking Status = ‘Current Smoker’ (VA Health Factor) |
| Diabetes mellitus | E10.x- E11.x (Type 1 and Type 2 diabetes) |
| Chronic heart disease | I25.x (Chronic ischemic heart disease), I50.x (Heart failure) |
| Chronic liver disease | K70.x (Alcoholic liver disease), K73.x-K74.x (Chronic hepatitis, fibrosis, cirrhosis) |
| Chronic lung disease | J43.x (Emphysema, unspecified), J44.x (COPD), J45.x (Asthma), J84.x (Interstitial lung disease) |

ICD-10: International Classification of Diseases, Tenth Revision, Clinical Modification;HIV: Human Immunodeficiency Virus; CSF: Cerebrospinal Fluid; COPD: Chronic Obstructive Pulmonary Disease; VA: Veterans Affairs

Supplemental Table 3a: Adjusted Variables in the Multivariable Logistic Regression Models

|  | **≥18 years stratified by Age groups** | | | | | | **≥65 years** | | | | | | | |
| --- | --- | --- | --- | --- | --- | --- | --- | --- | --- | --- | --- | --- | --- | --- |
| **List of Variables** | **Overall** | **18-49** | **50-64** | **65-74** | **75-84** | **85+** | **65+** | **High-risk** | **Medium-risk** | **Low-risk** | **West** | **Northeast** | **South** | **Midwest** |
| Age groups | X |  |  |  |  |  | X | X | X | X | X | X | X | X |
| Sex | X | X | X | X | X | S | X | X | X | X | X | X | X | X |
| Race | X | X | X | X | X | X | X | X | X | X | X | X | X | X |
| Ethnicity | X | X | X | X | S | S | X | S | S | S | S | S | X |  |
| **High-risk (IC)^$^** |  | X | X |  |  | X | X |  |  |  | X | X | X | X |
| Chronic renal failure | X |  |  | X | X |  |  | X |  |  |  |  |  |  |
| Generalized malignancy | X |  |  | X | X |  |  | X |  |  |  |  |  |  |
| Immunodeficiency | X |  |  | X | X |  |  | X |  |  |  |  |  |  |
| Iatrogenic immunosuppression | X |  |  | X | X |  |  | X |  |  |  |  |  |  |
| Leukemia | X |  |  | X | X |  |  | X |  |  |  |  |  |  |
| Lymphoma | X |  |  | X | X |  |  | X |  |  |  |  |  |  |
| Solid organ transplant | X |  |  | X | X |  |  | X |  |  |  |  |  |  |
| Other immunocompromised conditions^ⴕ^ | X |  |  | X | X |  |  | X |  |  |  |  |  |  |
| **Medium-risk (CMC) ^$^** |  |  |  |  |  | X |  |  |  |  |  |  |  |  |
| Chronic heart disease | X | X | X | X | X |  | X |  | X |  | X | X | X | X |
| Chronic lung disease | X | X | X | X | X |  | X |  | X |  | X | X | X | X |
| Chronic liver disease | X | X | X | X | X |  | X |  | X |  | X | X | X | X |
| Diabetes | X | X | X | X | X |  | X |  | X |  | X | X | X | X |
| Alcoholism | X | X | X | X | X |  | X |  | X |  | X | X | X | X |
| Smoking | X | X | X | X | X |  | X |  | X |  | X | X | X | X |
| Charlson Comorbidity Index score | X | S | S | X | X | X | X |  | X |  | X | X | X | X |
| Area Deprivation Index (ADI) | X | X | X | X | X | X | X | X | X | X | X | X | X | X |
| PPSV23 vaccination*^@^* | X | **S** | X | X | X | X | X | X | X | X | X | X | X | X |
| Influenza vaccination* | X | X | X | X | X | X | X | X | X | X | X | X | X | X |
| COVID-19 vaccination* | X | X | X | X | X | X | X | X | X | X | X | X | X | X |
| Index Year | X | X | X | X | X | X | X | X | X | X | X | X | X | X |
| Number of outpatient visits | X | X | X | X | X | X | X | S | X | X | X | X | X | X |
| Nursing home admission* | X | X | X | X | X | S | X | X | X | S | X | X | X | X |
| Intensive Care Unit admission* | X | X | X | X | X | X | X | X | X | S | X | X | X | X |
| Hospitalization* | X | X | X | X | X | X | X | X | X | X | X | X | X | X |
| Emergency department visit* | X | X | X | X | X | X | X | X | X | X | X | X | X | X |
| Region | X | X | X | X | X | X | X | X | X | X |  |  |  |  |

^ⴕ^Conditions with small cell count adjusted as a group variable (<2%; Cochlear, asplenia, cerebrospinal fluid (CSF) leak, HIV infection, Hodgkin, multiple myeloma, nephrotic syndrome, sickle cell disease and other hemoglobinopathies);

* Within prior 365 days from index date; ^@^ Within prior 5 years from index date; ^$^High- and medium-risk categories denote composite variables defined by ≥1 qualifying condition within each risk group and modeled as proxies for individual conditions due to small cell counts; PPSV23(23-valent pneumococcal polysaccharide vaccine); PCV13 (13-valent pneumococcal conjugate vaccine); COVID-19 (coronavirus disease 2019); IC: Immunocompromised Conditions; CMC: Chronic Medical Conditions; S: Eligible variable not included in the model due to extremely small cell count. X: Variable was included in the model

Supplemental Table 3b: Variables Included in the Propensity Score Models

|  | **≥18 years stratified by Age groups** | | | | | | **≥65 years** | | | | | | | |
| --- | --- | --- | --- | --- | --- | --- | --- | --- | --- | --- | --- | --- | --- | --- |
| **List of Variables** | **Overall** | **18-49** | **50-64** | **65-74** | **75-84** | **85+** | **65+** | **High-risk** | **Medium-risk** | **Low-risk** | **West** | **Northeast** | **South** | **Midwest** |
| Age groups | X |  |  |  |  |  | X | X | X | X | X | X | X | X |
| Sex | X | S | S | X | X | X | X | X | X | X | X | X | X | X |
| Race | X | X | X | X | X | X | X | X | X | X | X | X | X | X |
| Ethnicity | X | X | X | X | S | X | X | S | S |  | S | S | X | X |
| **High-risk (IC)** ^$^ |  | X | X |  |  | X | X |  |  |  | X | X | X | X |
| Chronic renal failure | X |  |  | X | X |  |  | X |  |  |  |  |  |  |
| Generalized malignancy | X |  |  | X | X |  |  | X |  |  |  |  |  |  |
| Immunodeficiency | X |  |  | X | X |  |  | X |  |  |  |  |  |  |
| Iatrogenic immunosuppression | X |  |  | X | X |  |  | X |  |  |  |  |  |  |
| Leukemia | X |  |  | X | X |  |  | X |  |  |  |  |  |  |
| Lymphoma | X |  |  | X | X |  |  | X |  |  |  |  |  |  |
| Solid organ transplant | X |  |  | X | X |  |  | X |  |  |  |  |  |  |
| Other immunocompromised conditions^ⴕ^ | X |  |  | X | X |  |  | X |  |  |  |  |  |  |
| **Medium-risk (CMC)** ^$^ |  | X | X |  |  | X |  |  |  |  |  |  |  |  |
| Chronic heart disease | X |  |  | X | X |  | X |  | X |  | X | X | X | X |
| Chronic lung disease | X |  |  | X | X |  | X |  | X |  | X | X | X | X |
| Chronic liver disease | X |  |  | X | X |  | X |  | X |  | X | X | X | X |
| Diabetes | X |  |  | X | X |  | X |  | X |  | X | X | X | X |
| Alcoholism | X |  |  | X | X |  | X |  | X |  | X | X | X | X |
| Smoking | X |  |  | X | X |  | X |  | X |  | X | X | X | X |
| Charlson Comorbidity Index score | X | S | S | X | X | X | X | S | X |  | X | X | X | X |
| Area Deprivation Index (ADI) | X | X | X | X | X | X | X | X | X | X | X | X | X | X |
| PPSV23 vaccination*^@^* | X | S | X | X | X | X | X | X | X | X | X | X | X | X |
| Influenza vaccination* | X | X | X | X | X | X | X | X | X | X | X | X | X | X |
| COVID-19 vaccination* | X | X | X | X | X | X | X | X | X | X | X | X | X | X |
| Index Year | X | X | X | X | X | X | X | X | X | X | X | X | X | X |
| Number of outpatient visits | X | X | X | X | S | X | X | S | X | S | X | X | X | X |
| Nursing home admission* | X | X | X | X | X | S | X | X | X | S | X | X | X | X |
| Intensive Care Unit admission* | X | X | X | X | X | X | X | X | X | X | X | X | X | X |
| Hospitalization* | X | X | X | X | X | X | X | X | X | X | X | X | X | X |
| Emergency department visit* | X | X | X | X | X | X |  | X | X | X | X | X | X | X |
| Region | X | X | X | X | X | X |  | X | X | X |  |  |  |  |

^ⴕ^Conditions with small cell count adjusted as a group variable (<2%; Cochlear, asplenia, cerebrospinal fluid (CSF) leak, HIV infection, Hodgkin, multiple myeloma, nephrotic syndrome, sickle cell disease and other hemoglobinopathies);

* Within prior 365 days from index date; ^@^ Within prior 5 years from index date; ^$^High- and medium-risk categories denote composite variables defined by the presence of ≥1 qualifying condition within each risk group and were used in subgroup analyses due to small cell counts; PPSV23(23-valent pneumococcal polysaccharide vaccine); PCV13 (13-valent pneumococcal conjugate vaccine); COVID-19 (coronavirus disease 2019); IC: Immunocompromised Conditions; CMC: Chronic Medical Conditions; S: Eligible variable not included in the model due to extremely small cell count. X: Variable was included in the model.

Supplemental Table 4: Sensitivity analysis of PCV20 effectiveness against pneumococcal pneumonia hospitalization allowing previous PCV13 vaccination, January 2023-December 2025 (N=36,641)

|  |  | **Unadjusted VE (95% CI)** | **Multivariable (95% CI)** | **IPTW (95% CI)** |
| --- | --- | --- | --- | --- |
| **Age groups** | 18+ years (36641) | -0.51 (-18.22, 14.55) | 6.97 (-10.46, 21.65) | 6.83 (-9.85, 20.98) |
|  | 18-49 years (984) | 61.51 (-67.16, 91.14) | 40.10 (-192.74, 87.74) | 40.43 (-121.84, 84.00) |
|  | 50-64 years (4535) | **-55.88 (-121.75, -9.58)** | **-58.23 (-133.12, -7.40)** | **-52.99 (-115.99, -8.37)** |
|  | 65-74 years (11358) | 9.14 (-17.75, 29.88) | 10.66 (-17.65, 32.17) | -0.66 (-28.84, 21.36) |
|  | 75-84 years (15382) | 15.12 (-16.09, 37.94) | 17.57 (-13.61, 40.19) | 13.97 (-17.49, 37.00) |
|  | 85+ years (4382) | 52.20 (-10.62, 79.35) | 50.55 (-16.09, 78.93) | 52.96 (-9.30, 79.75) |
| **≥65 years** | 65 years and above (31122) | 11.15 (-7.59, 26.63) | 18.03 (-0.17, 32.92) | 16.24 (-1.62, 30.96) |
|  | **Risk group** |  |  |  |
|  | High risk (14391) | 1.59 (-32.99, 27.18) | 7.71 (-26.55, 32.69) | 9.70 (-22.80, 33.60) |
|  | Medium risk (15859) | 17.87 (-5.81, 36.25) | **23.71 (0.49, 41.52)** | 21.87 (-0.92, 39.51) |
|  | Low risk (872) | 10.94 (-205.34, 74.02) | 10.67 (-252.54, 77.37) | 49.40 (-123.49, 88.54) |
|  | **U.S. census regions** |  |  |  |
|  | West (6370) | 24.85 (-15.89, 51.27) | 35.44 (-1.71, 59.03) | **46.65 (13.41, 67.13)** |
|  | Northeast (4548) | 4.78 (-46.48, 38.11) | 4.34 (-50.50, 39.20) | 8.71 (-41.68, 41.18) |
|  | South (13662) | -0.42 (-41.12, 28.54) | 4.89 (-35.87, 33.42) | -2.50 (-43.06, 26.56) |
|  | Midwest (6542) | 10.83 (-27.49, 37.63) | 25.83 (-8.03, 49.07) | 21.39 (-13.71, 45.66) |

Bold indicates statistically significant, >0 indicates vaccine effectiveness, <0 indicates vaccine ineffectiveness.

Supplemental Table 5: Negative Control Analysis for IPTW Analysis in the Primary Analysis

|  |  | **Fracture** | **Cataract** |
| --- | --- | --- | --- |
| **Age groups** | 18 and above (34009) | -0.33 (-11.72, 9.89) | 3.20 (-3.14, 9.15) |
|  | 18-49 years (946) | -29.66 (-196.22, 43.24) | -57.39 (-409.00, 51.33) |
|  | 50-64 years (4203) | -6.54 (-41.78, 19.94) | -7.54 (-27.95, 9.62) |
|  | 65-74 years (10140) | 8.25 (-9.15, 22.88) | -1.40 (-11.24, 7.56) |
|  | 75-84 years (14576) | -0.21 (-19.39, 15.90) | 9.70 (-0.16, 18.60) |
|  | 85+ years (4144) | -26.90 (-69.35, 4.90) | 14.44 (-10.11, 33.52) |
| **≥65 years** | 65 and above (28860) | -0.65 (-12.52, 9.96) | 6.02 (-0.36, 11.99) |
|  | **Risk group** |  |  |
|  | High-risk (IC) (13318) | 3.30 (-13.59, 17.67) | 3.33 (-6.49, 12.25) |
|  | Medium-risk (CMC) (14725) | -9.63 (-27.95, 6.06) | **-9.47 (-19.72, -0.09)** |
|  | Low risk (817) | 19.96 (-115.90, 70.33) | 12.16 (-45.47, 46.96) |
|  | **U.S. Regions** |  |  |
|  | West (5886) | 11.35 (-12.34, 30.04) | 0.77 (-14.92, 14.31) |
|  | Northeast (4303) | -8.59 (-46.07, 19.27) | 8.70 (-7.64, 22.56) |
|  | South (12618) | -7.45 (-26.78, 8.93) | **-14.25 (-26.00, -3.60)** |
|  | Midwest (6053) | -1.54 (-31.22, 21.43) | -7.00 (-23.52, 7.31) |

IPTW, inverse probability of treatment weighted; IC: Immunocompromised Conditions; CMC: Chronic Medical Conditions (CMC)

Cataract (ICD-10-CM H25.x).

Fracture due to trauma (ICD-10-CM S22.x, S32.x, S42.x, S52.x, S62.x, S82.x, S92.x).

Bold indicates statistically significant, >0 indicates vaccine effectiveness, <0 indicates vaccine ineffectiveness.

**REFERENCES:**

1.	Smithee, R. B. *et al.* Pneumonia Hospitalization Coding Changes Associated With Transition From the 9th to 10th Revision of International Classification of Diseases. *Health Serv Res Manag Epidemiol* **7**, 2333392820939801 (2020).

2.	Kobayashi, M. Expanded Recommendations for Use of Pneumococcal Conjugate Vaccines Among Adults Aged ≥50 Years: Recommendations of the Advisory Committee on Immunization Practices — United States, 2024. *MMWR Morb Mortal Wkly Rep* **74**, (2025).
